# A Behaviorally Informed Patient-Facing Intervention to Promote Albuminuria Testing in Hypertension: A Mixed-Methods Evaluation of KnowCKD

**DOI:** 10.64898/2026.11.06.26367454

**Authors:** Ivan Villasenor, Rahul Biswas, Kelechi Okpara, Sharlene Shirali, Susanne B. Nicholas, Jiun-Ruey Hu, Delphine S. Tuot, Nicole A. Errett, Julio A. Lamprea-Montealegre

## Abstract

**Background:** Albuminuria testing to diagnose chronic kidney disease (CKD) and guide blood pressure management is recommended for adults with hypertension but remains underused in part due to suboptimal patient understanding of its importance. Patient-facing behavioral interventions have the potential to bridge this gap. We developed and evaluated KnowCKD, a behaviorally informed intervention designed to increase patient awareness of CKD and knowledge and promote albuminuria testing among adults with hypertension.

**Methods:** KnowCKD consisted of an educational infographic and brief, behaviorally informed messages delivered to patients’ smartphones during the 5 days leading up to and including a scheduled primary care visit at one academic medical center. Among 57 adults with hypertension without diabetes or recent albuminuria testing, we assessed changes in CKD awareness and knowledge and patient-reported engagement via survey pre- and post-exposure to KnowCKD. Electronic Health Record (EHR)-confirmed albuminuria or proteinuria testing was compared with 182 contemporaneous primary care controls using unmatched and propensity score-matched analysis. A post-intervention focus group evaluated participant experiences.

**Results:** After exposure to KnowCKD, recognition that adults with hypertension should undergo urine testing increased from 58% to 95%, knowledge that albuminuria indicates protein in the urine increased from 47% to 95%, and awareness that testing facilitates early CKD detection increased from 55% to 100% (P <0.001 for all). After the primary care visit, 77% of patients reported asking their clinician about urine testing and 65% reported that testing was ordered. EHR-confirmed albuminuria or proteinuria testing occurred in 36.8% of KnowCKD participants versus 11.0% of controls (risk ratio, 3.35 [95% CI, 1.96–5.73]; absolute risk difference, 25.9 percentage points [95% CI, 12.5–39.2]). The propensity score–matched risk ratio was 2.93 (95% CI, 1.59–5.42). Participants described KnowCKD as timely, understandable, and actionable and emphasized trusted health-system communication for broader implementation.

**Conclusions:** KnowCKD was associated with increased patient awareness of CKD and knowledge, patient engagement, and higher EHR-confirmed albuminuria or proteinuria testing among adults with hypertension. A rigorous evaluation of KnowCKD implementation within routine health-system workflows is warranted as a next step.

## Introduction

Hypertension is a major cause of chronic kidney disease (CKD), a condition associated with substantially increased cardiovascular morbidity and mortality. Approximately 1 in 5 adults with hypertension without diabetes have CKD,^1,2^ defined by an estimated glomerular filtration rate (eGFR) below 60 mL/min/1.73 m² and/or albuminuria, defined as a urine albumin-to-creatinine ratio (UACR) ≥30 mg/g. Among adults with hypertension, detection of albuminuria is critical for cardiovascular and kidney risk stratification and for identifying patients who may benefit from therapies such as angiotensin-converting enzyme inhibitors, angiotensin receptor blockers, and sodium-glucose cotransporter-2 inhibitors, which reduce cardiovascular and kidney risk. Accordingly, the 2025 ACC/AHA hypertension guideline recommends UACR measurement as part of the routine evaluation of hypertension.^3^ This is consistent with the 2023 American Heart Association cardiovascular-kidney-metabolic (CKM) syndrome Presidential Advisory.^4^

Despite its clinical importance, albuminuria testing remains underused. Approximately 5% of adults with hypertension without diabetes undergo albuminuria testing, leaving nearly 90% of prevalent albuminuria undetected.^5^ Although routine UACR testing in hypertension has only recently been incorporated in major hypertension guidelines, low testing rates also likely reflect longstanding known barriers to CKD detection and care, including limited patient awarenss of CKD and the role of albuminuria testing.^6^ These barriers operate across patient, clinician, and health-system levels.^7^ Efforts to improve CKD detection have frequently focused on clinician- and health-system–level strategies, with comparatively less attention to engaging patients directly in the detection process.^8–10^ Patient-facing interventions may provide a complementary approach by increasing CKD awareness and knowledge while enabling patients to initiate discussions about guideline-recommended testing during clinical encounters.

Patient activation strategies informed by behavioral science and behavioral economics offer a promising approach to addressing persistent gaps in evidence-based care.^11,12^ By increasing the salience of recommended actions, reducing barriers to action, and prompting patients to engage clinicians around specific care opportunities, these interventions may simultaneously improve patients’ awareness and knowledge while promoting action during clinical encounters. Behaviorally informed nudges have improved uptake of preventive services, including vaccination, and patient-facing interventions delivered before clinical encounters have also influenced treatment decisions.^11,13,14^ In the EPIC-HF randomized trial for example, an electronically delivered patient-activation tool provided before cardiology visits facilitated clinician intensification of guideline-directed medical therapy among patients with heart failure.^15^

Building on these principles, we developed KnowCKD, a patient-facing, behaviorally informed intervention consisting of an educational infographic and brief messages delivered before a scheduled primary care visit. KnowCKD was designed to increase CKD awareness and knowledge and encourage patients to discuss albuminuria testing with their clinicians. In this mixed-methods study, we assessed changes in CKD awareness and knowledge and patient engagement compared electronic health record– confirmed albuminuria or proteinuria testing between KnowCKD participants and contemporaneous primary care controls, and evaluated participants’ experiences and considerations for implementation within routine clinical care.

## Methods

### Study Design and Setting

KnowCKD was developed and evaluated through a sequential study involving patient-informed intervention development guided by behavioral science principles, implementation in clinical care, and a mixed-methods evaluation. The evaluation included: (1) prospective assessment of changes in CKD awareness and knowledge and patient-reported engagent, defined by discussion of albuminuria testing with the primary care clinician; (2) an observational comparison of electronic health record (EHR)-confirmed albuminuria or proteinuria testing between KnowCKD participants and contemporaneous primary care controls; and (3) a post-intervention qualitative evaluation examining participants’ experiences with the intervention and their considerations for broader implementation.

The study was conducted at one academic medical center between August 2025 and March 2026. The study was approved by the University of California San Francisco (UCSF) Institutional Review Board [24-41224], and KnowCKD participants provided written informed consent.

### Development of KnowCKD

KnowCKD was developed using a patient-informed, human-centered design process guided by behavioral science and behavioral economics and consistent with established methods for the iterative development of health-related messages.^11,16,17^ Formative development involved 10 English-speaking adults with self-reported hypertension, with or without CKD, recruited from the Health eHeart Study through the Eureka digital research platform.^18^ Patitents were required to by English speadking. No additional inclusion or exclusion criteria were applied

Participants took part in 3 sequential video-based discussion sessions and provided feedback on message content, terminology, communication strategies, and questionnaire design. Feedback was used iteratively to refine the intervention.

The intervention delivered to participants consisted of an educational infographic followed by a series of brief, behaviorally informed messages delivered before a scheduled primary care visit. Content was organized around 5 behavioral principles: salience and awareness, simplification and friction reduction, loss aversion and risk framing, patient activation and self-efficacy, and timely prompting (**Table 1**).

**Table 1.**
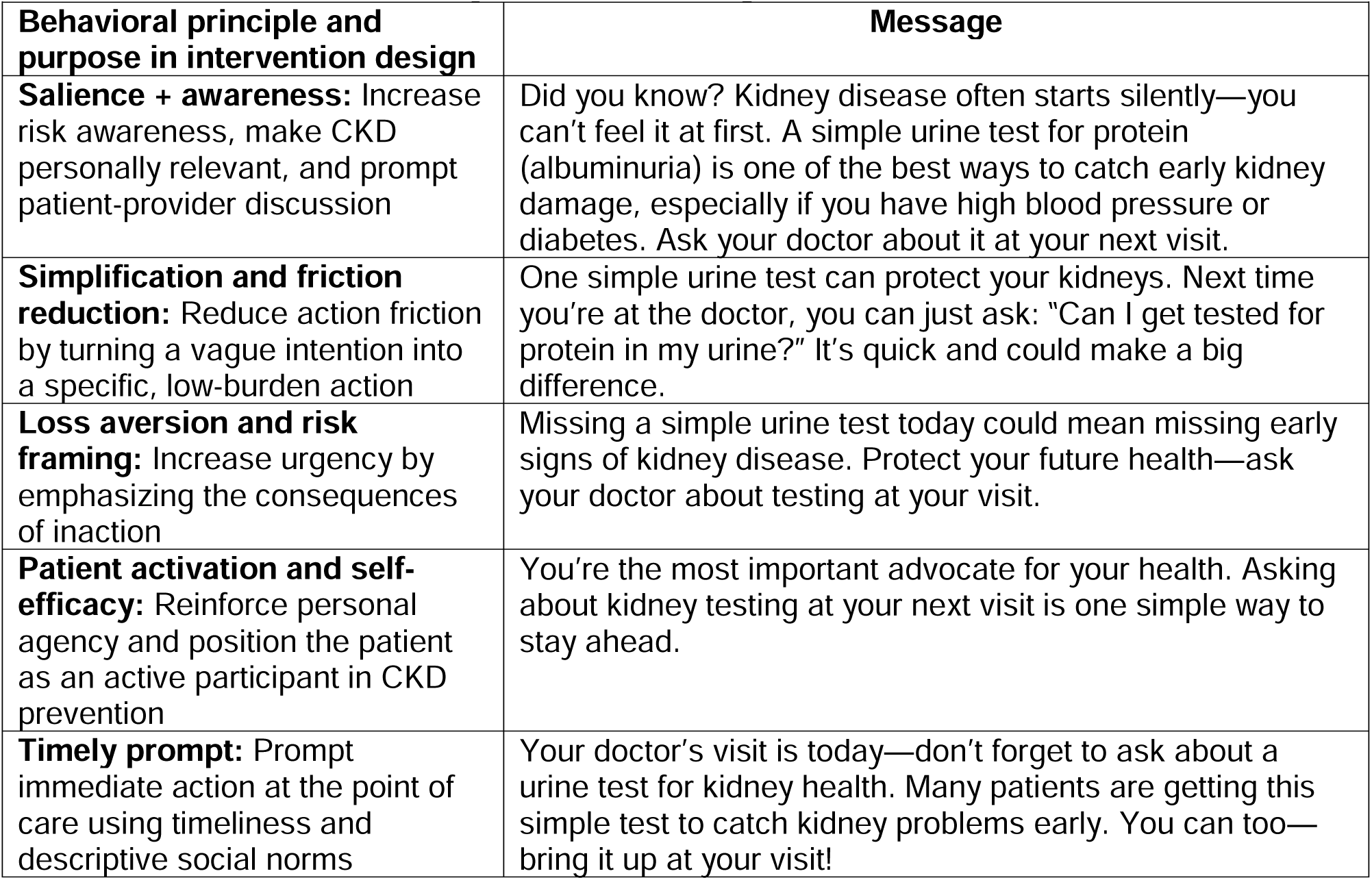
KnowCKD behaviorally informed messages.

| **Behavioral principle and purpose in intervention design** | **Message** |
|---|---|
| **Salience + awareness:** Increase risk awareness, make CKD personally relevant, and prompt patient-provider discussion | Did you know? Kidney disease often starts silently—you can’t feel it at first. A simple urine test for protein (albuminuria) is one of the best ways to catch early kidney damage, especially if you have high blood pressure or diabetes. Ask your doctor about it at your next visit. |
| **Simplification and friction reduction:** Reduce action friction by turning a vague intention into a specific, low-burden action | One simple urine test can protect your kidneys. Next time you’re at the doctor, you can just ask: “Can I get tested for protein in my urine?” It’s quick and could make a big difference. |
| **Loss aversion and risk framing:** Increase urgency by emphasizing the consequences of inaction | Missing a simple urine test today could mean missing early signs of kidney disease. Protect your future health—ask your doctor about testing at your visit. |
| **Patient activation and self-efficacy:** Reinforce personal agency and position the patient as an active participant in CKD prevention | You’re the most important advocate for your health. Asking about kidney testing at your next visit is one simple way to stay ahead. |
| **Timely prompt:** Prompt immediate action at the point of care using timeliness and descriptive social norms | Your doctor’s visit is today—don’t forget to ask about a urine test for kidney health. Many patients are getting this simple test to catch kidney problems early. You can too—bring it up at your visit! |

Messages emphasized that early kidney disease may be asymptomatic, introduced urine testing for protein as a means of detecting early kidney damage, provided simple language patients could use to discuss testing with their clinician, described potential consequences of missed detection, reinforced patients’ role as active participants in their care, and included a reminder to request albuminuria testing on the day of the scheduled visit.

### Study Population and Recruitment

Participants were adults receiving primary care at a single academic medical center with documented hypertension, without diabetes, and without albuminuria testing during the preceding 12 months. Eligible participants were also required to use MyChart, have access to a smartphone, and have an upcoming primary care appointment scheduled at least 1 week after enrollment, allowing sufficient time for delivery of KnowCKD before the clinical encounter.

Potentially eligible participants were identified through a UCSF EHR-based recruitment service that conducted daily searches of primary care schedules and sent electronic invitations to patients meeting initial study criteria. A total of 406 patients expressed preliminary interest through this recruitment process. Among these, 84 completed the MyChart-based study sign-up process and met the additional requirement of having an upcoming primary care appointment within the specified enrollment window. These patients were contacted by the study team to confirm eligibility and obtain written informed consent, and 57 ultimately enrolled in KnowCKD.

Patients identified through the same EHR-based recruitment process who did not enroll in KnowCKD constituted the pool of potential controls for the comparative analysis. Following enrollment, KnowCKD participants completed the pre-intervention questionnaire and received the intervention through the Eureka digital research platform. Messages were delivered to participants’ smartphones at fixed intervals during the 5 days leading up to and including their scheduled primary care visit. The date of completion of the pre-intervention questionnaire served as the index date and marked the beginning of exposure to KnowCKD.

### Survey Measures

Participants completed a questionnaire before receiving KnowCKD and a follow-up questionnaire after their primary care visit. Surveys assessed CKD awareness and knowledge and patient-reported engagement related to albuminuria testing.

The awareness and knowledge items were developed by the study team for this intervention and were not drawn from a previously validated questionnaire. Items assessed whether participants recognized that early CKD may occur without symptoms, that adults with hypertension should undergo urine testing for kidney disease, that albuminuria refers to protein in the urine, and that albuminuria testing can facilitate early CKD detection. The wording and content of these items were refined based on feedback from the 10 participants who took part in the formative development discussions.

Post-intervention survey items also assessed whether participants reported discussing urine testing with their clinician and whether their clinician ordered a urine test. These patient-reported measures were analyzed separately from the EHR-confirmed testing outcome.

### EHR-Confirmed Albuminuria or Proteinuria Testing

The EHR-confirmed outcome was completion of albuminuria or proteinuria testing, defined as documentation in the EHR of either UACR or urine protein-to-creatinine ratio (UPCR), identified using procedure codes.

For KnowCKD participants, the index date was the date of completion of the pre-intervention questionnaire. Follow-up began on this date and continued through the data extraction date of April 30, 2026. Any qualifying test completed during this period was considered post-intervention testing. Follow-up varied according to enrollment date, with a median observation period of 188 days.

From the pool of potential controls described above, we selected contemporaneous controls whose most recent primary care visit occurred during the KnowCKD enrollment window (August 2025 through March 2026). This restriction yielded 182 controls. For controls, albuminuria or proteinuria testing was assessed during the 12 months preceding the data extraction date of April 30, 2026, providing an estimate of annual testing during the study period.

Because absence of albuminuria or proteinuria testing during the preceding 12 months was an intended eligibility criterion, we conducted a sensitivity analysis excluding patients with qualifying testing during that period. EHR review identified prior testing in 2 KnowCKD participants and 18 controls. The primary and propensity score–matched analyses were repeated after excluding these patients.

The resulting primary analytic cohort included 57 KnowCKD participants and 182 contemporaneous controls.

### Qualitative Evaluation

Following exposure to KnowCKD and the associated primary care visit, 8 participants took part in a single 60-minute facilitated focus group. To minimize recall bias, recruitment was restricted to participants enrolled in KnowCKD during 2026. Participants were recruited from survey respondents who had indicated willingness to be contacted for further research. They were purposively selected to include individuals who had and had not completed albuminuria testing, allowing the focus group to capture experiences across different post-intervention testing outcomes.

The semi-structured discussion guide was informed by the Consolidated Framework for Implementation Research (CFIR) and explored participants’ recall of and reactions to the intervention,^19^ impressions of the messages and infographic, interactions with primary care clinicians, whether albuminuria testing was discussed, ordered, or completed, subsequent actions, message volume and timing, terminology, trust in communication channels, and practical barriers to completing testing.

We used a hybrid deductive-inductive thematic analysis. Deductive coding was informed by CFIR constructs relevant to participants’ experiences with the intervention, while inductive coding captured concepts emerging from the focus group, including patient engagement, message timing, trust in the communication source, terminology, clinician receptivity, and practical barriers to test completion. Coding was performed by a single investigator. Codes were iteratively reviewed and grouped into higher-order categories, which were refined into themes representing recurrent patterns in participants’ experiences and perceptions of KnowCKD.

### Statistical Analysis

For survey outcomes, within-participant changes in binary measures of CKD awareness and knowledge between pre- and post-intervention assessments were evaluated using exact McNemar tests. Post-intervention proportions of participants reporting that they discussed urine testing with their clinician and that their clinician ordered a urine test were summarized descriptively.

For the primary EHR-based analysis, we compared the proportion of patients completing albuminuria or proteinuria testing between KnowCKD participants and contemporaneous controls using the χ² test. Risk ratios and corresponding 95% confidence intervals were calculated using the Katz log method. Absolute risk differences and their 95% confidence intervals were calculated using the Wald method.

Contemporaneous controls were defined as patients with hypertension whose primary care visit fell within the KnowCKD enrollment window (August 2025 to March 2026) and who did not participate in KnowCKD. Propensity scores were estimated using logistic regression with KnowCKD participation as the dependent variable. The model included 8 prespecified baseline characteristics: age, sex, race and ethnicity, number of primary care visits during the preceding 3 years, most recently recorded systolic blood pressure, number of antihypertensive medication classes, most recently recorded eGFR, and use of an angiotensin-converting enzyme inhibitor or angiotensin receptor blocker.

KnowCKD participants were matched to up to 2 contemporaneous controls using greedy nearest-neighbor matching without replacement, with a caliper of 0.2 times the standard deviation of the logit propensity score. Four KnowCKD participants could not be matched within the caliper and were excluded from the matched cohort. The standardized mean difference (SMD) before matching compared all KnowCKD participants (n=57) with all contemporaneous controls (n=182); the SMD after matching compared matched KnowCKD participants (n=53) with matched controls (n=101). An absolute SMD below 0.10 was considered indicative of adequate balance. In the matched cohort, risk ratios and absolute risk differences were estimated using the same methods as in the primary analysis, without additional regression adjustment.

In a sensitivity analysis, although KnowCKD eligibility criteria required no albuminuria or proteinuria testing during the preceding 12 months, verification of laboratory records identified 2 KnowCKD participants with qualifying testing before or at enrollment. To assess the potential influence of prior testing, we repeated the primary and propensity score–matched analyses after excluding KnowCKD participants and controls with documented UACR or UPCR testing during the preceding 12 months. For the propensity score–matched analysis, exclusions were applied to the original matched cohort without repeating the matching procedure.

All statistical tests were 2-sided, and P<0.05 was considered statistically significant. Analyses were performed using R version 4.4 (R Foundation for Statistical Computing, Vienna, Austria).

## Results

Among 406 patients who expressed preliminary interest through the EHR-based recruitment process, 84 completed the MyChart-based study sign-up process and met the additional appointment-related eligibility requirement. Of these, 57 enrolled in KnowCKD and completed the pre-intervention assessment. Based on the pre-intervention questionnaire, participants had a mean age of 60.0 years, 54.4% were female, and 57.9% identified as White. Two participants (3.5%) reported having CKD, while 4 (7.0%) were uncertain of their CKD status (**Table 2**).

**Table 2.** Demographic and clinical characteristics of KnowCKD participants (N = 57). Characteristic N.

| **Characteristic** | **N = 57** |
|---|---|
| **Age (years)** |  |
| Mean (SD) | 60.0 (12.6) |
| Median [IQR] | 60 [52–70] |
| Missing | 3 (5.3%) |
| **Sex, n (%)** |  |
| Female | 31 (54.4%) |
| Male | 26 (45.6%) |
| **Race, n (%)<sup>1</sup>** |  |
| White | 33 (57.9%) |
| Asian | 14 (24.6%) |
| Black or African American | 7 (12.3%) |
| Native Hawaiian or Other Pacific Islander | 2 (3.5%) |
| American Indian or Alaska Native | 0 (0.0%) |
| Other | 4 (7.0%) |
| **Ethnicity, n (%)** |  |
| Hispanic or Latino | 5 (8.8%) |
| Non-Hispanic or Latino | 51 (89.5%) |
| Missing | 1 (1.8%) |
| **Self-reported CKD status, n (%)** |  |
| Yes | 2 (3.5%) |
| No | 51 (89.5%) |
| Don’t know | 4 (7.0%) |

*Inclusion criteria:* adult primary care patients at UCSF with documented hypertension, no diabetes, and no albuminuria testing in the prior 12 months.
<sup>1</sup> Demographic characteristics and CKD status in this table were obtained from the pre-intervention questionnaire and are self-reported. Age was missing for 3 participants. Race was collected using a “select all that apply” format; therefore, categories are not mutually exclusive and percentages may sum to more than 100%. These self-reported characteristics may differ from EHR-derived variables used in the comparative analyses presented in Table 3.
All characteristics are self-reported via the pre-message survey. Abbreviations: CKD, chronic kidney disease; IQR, interquartile range; SD, standard deviation.

### CKD Awareness, Knowledge, and Patient Activation

CKD awareness and knowledge increased following exposure to KnowCKD (**Figure 1A**). The proportion of participants recognizing that adults with hypertension should undergo urine testing for kidney disease increased from 58% before the intervention to 95% afterward (P<0.001). Recognition that albuminuria refers to protein in the urine increased from 47% to 95% (P<0.001), while awareness that albuminuria testing facilitates early CKD detection increased from 55% to 100% (P<0.001). Knowledge that early CKD may occur without symptoms increased from 54% to 72%, although this difference did not reach statistical significance.

**Figure 1.**
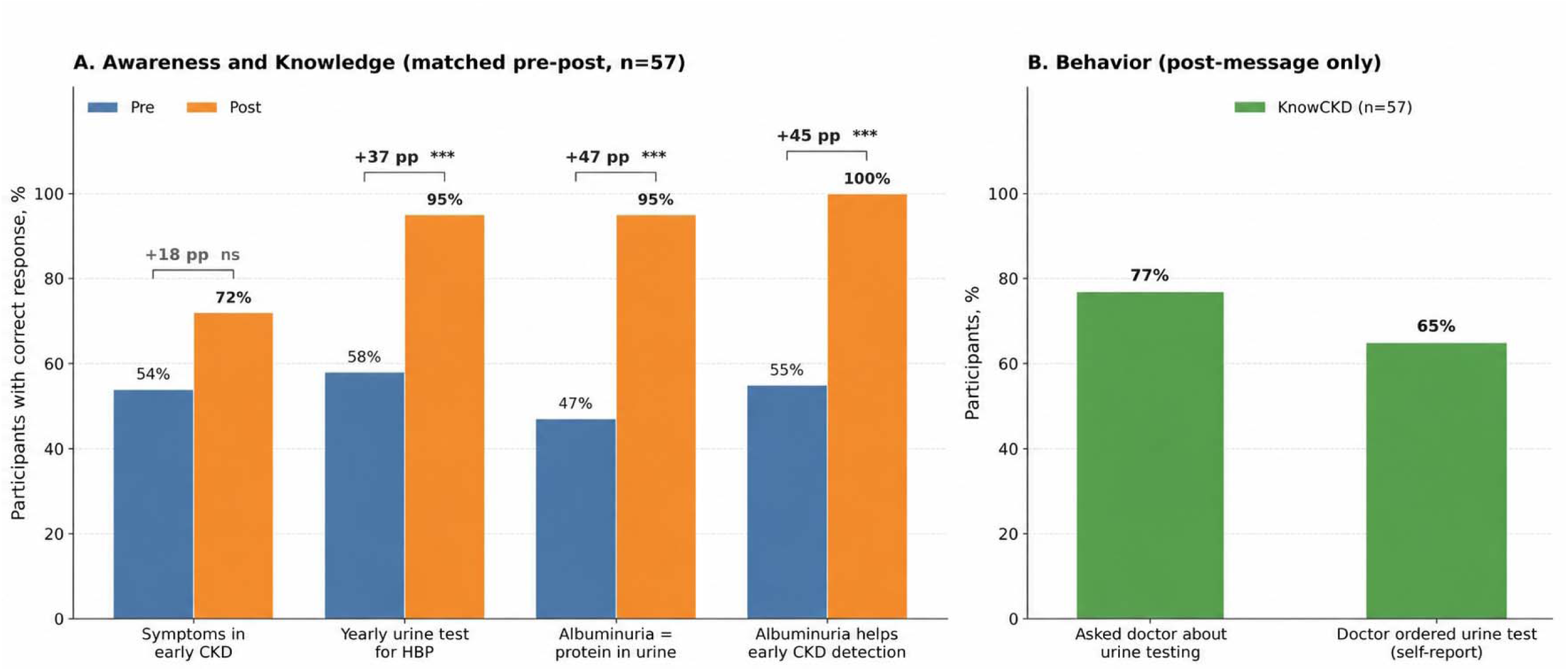
Changes in CKD Awareness and Knowledge and Self-Reported Patient engagement Following KnowCKD. **A:** Proportion of KnowCKD participants providing correct responses to four CKD awareness and knowledge questions before and after the intervention. Within-participant changes were evaluated using exact McNemar tests. **Panel B:** Proportion of KnowCKD participants reporting that they discussed urine testing with their clinician (77%) and that their clinician ordered a urine test (65%) following the intervention. These outcomes were self-reported and were not independently verified through electronic health records. **Abbreviations:** CKD, chronic kidney disease; HBP, high blood pressure; pp, percentage points. ***P<0.001; ns, not statistically significant.

KnowCKD was also associated with high levels of self-reported patient engagement (**Figure 1B**). Following their primary care visit, 77% of participants reported discussing urine testing with their clinician, and 65% reported that their clinician had ordered a urine test.

### EHR-Confirmed Albuminuria or Proteinuria Testing

During post-intervention follow-up, 21 of 57 KnowCKD participants (36.8%) completed UACR or UPCR testing, compared with 20 of 182 contemporaneous controls (11.0%). This corresponded to a risk ratio of 3.35 (95% CI, 1.96–5.73; P<0.001) and an absolute risk difference of 25.9 percentage points (95% CI, 12.5–39.2).

Propensity score matching was performed to improve comparability between KnowCKD participants and contemporaneous controls (**Table 3**; **Figure 2**). Before matching, the largest differences were observed in angiotensin-converting enzyme inhibitor or angiotensin receptor blocker use (50.9% vs 34.1%; standardized mean difference [SMD], 0.345), number of antihypertensive medication classes (1.2 vs 1.0; SMD, 0.257), and systolic blood pressure (126.7 vs 129.5 mm Hg; SMD, −0.202).

**Figure 2.**
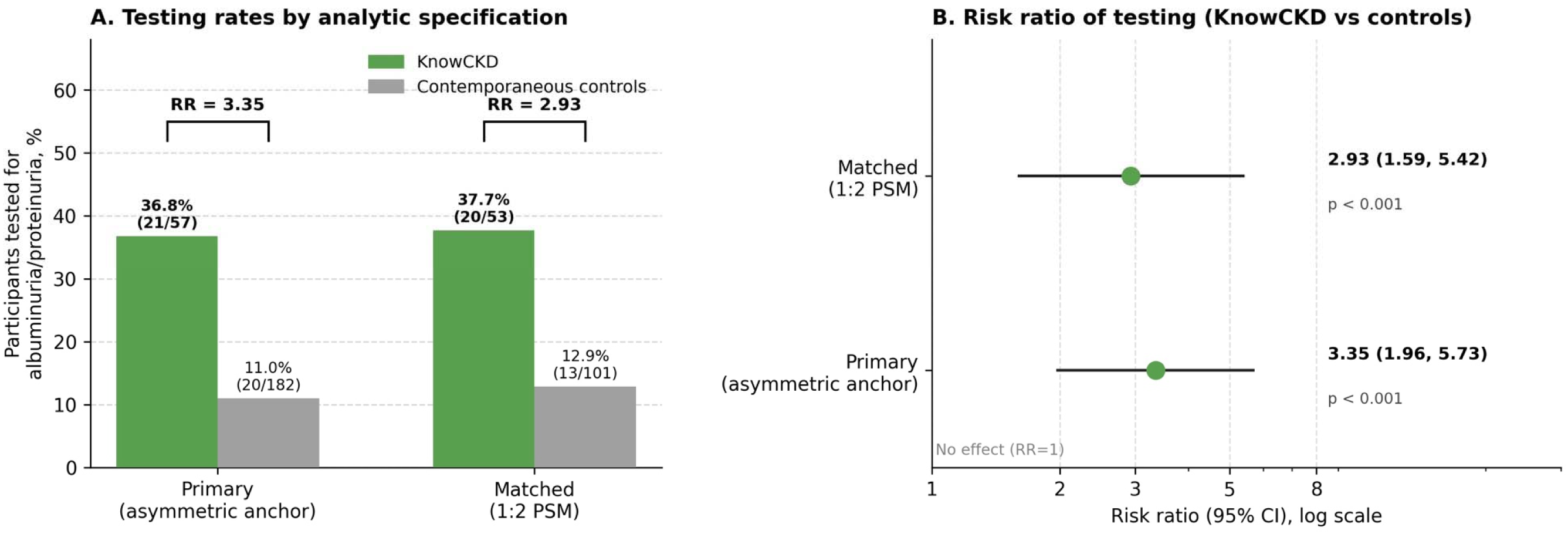
Association Between KnowCKD and EHR-Confirmed Albuminuria or Proteinuria Testing. **Panel A:** Proportion of KnowCKD participants and contemporaneous controls completing UACR or UPCR testing across three analytic specifications: the primary analysis, a sensitivity analysis applying the same 12-month calendar-based measurement window to both groups, and the 1:2 propensity score–matched analysis. **Panel B:** Corresponding risk ratios and 95% confidence intervals for each analysis. In the primary analysis, testing was assessed from enrollment through April 30, 2026, for KnowCKD participants and during the 12 months ending April 30, 2026, for controls. The propensity score–matched analysis included 53 KnowCKD participants and 101 controls. The dashed vertical line indicates a risk ratio of 1. **Abbreviations:** EHR, electronic health record; UACR, urine albumin-to-creatinine ratio; UPCR, urine protein-to-creatinine ratio.

**Table 3.** Baseline characteristics of KnowCKD participants and contemporaneous controls before and after 1:2 propensity score matching.

| Characteristic | KnowCKD (n=57) | All controls (n=182) | Matched KnowCKD (n=53) | Matched controls (n=101) | SMD before | SMD after |
|---|---|---|---|---|---|---|
| **Demographics** |  |  |  |  |  |  |
| Age (years), mean (SD) | 61.1 (12.6) | 62.8 (13.7) | 62.0 (12.6) | 62.4 (13.6) | -0.129 | -0.033 |
| Female, n (%) | 31 (54.4%) | 94 (51.6%) | 29 (54.7%) | 58 (57.6%) | +0.055 | -0.057 |
| Race: White, n (%) | 29 (50.9%) | 110 (60.4%) | 29 (54.7%) | 54 (53.8%) | -0.193 | +0.019 |
| Race: Asian, n (%) | 13 (22.8%) | 34 (18.7%) | 13 (24.5%) | 22 (21.7%) | +0.102 | +0.068 |
| Race: Black or African American, n (%) | 6 (10.5%) | 10 (5.5%) | 3 (5.7%) | 10 (10.4%) | +0.186 | -0.154 |
| Race: Latinx, n (%) | 3 (5.3%) | 15 (8.2%) | 3 (5.7%) | 4 (3.8%) | -0.119 | +0.085 |
| Race: Other/Multi/Unknown, n (%) | 6 (10.5%) | 13 (7.1%) | 5 (9.4%) | 10 (10.4%) | +0.119 | -0.031 |
| **Healthcare engagement** |  |  |  |  |  |  |
| PCP visits in past 3 years, mean (SD) | 8.9 (5.8) | 8.4 (5.9) | 8.6 (5.7) | 8.8 (6.5) | +0.099 | -0.037 |
| **Hypertension and kidney profile** |  |  |  |  |  |  |
| Years with hypertension, median [IQR] | 9 [3–13] | 9 [4–13] | 9 [3–13] | 9 [4–13] |  |  |
| Systolic blood pressure (mmHg), mean (SD) | 126.7 (14.1) | 129.5 (14.2) | 126.6 (14.1) | 127.5 (14.0) | -0.202 | -0.064 |
| # BP medication classes, mean (SD) | 1.2 (0.9) | 1.0 (0.9) | 1.2 (0.9) | 1.1 (0.9) | +0.257 | +0.043 |
| eGFR (mL/min/1.73 m²), mean (SD) | 86.2 (17.6) | 83.6 (18.7) | 84.7 (17.4) | 83.8 (17.0) | +0.140 | +0.048 |
| On ACEi or ARB, n (%) | 29 (50.9%) | 62 (34.1%) | 25 (47.2%) | 43 (42.4%) | +0.345 | +0.094 |

Contemporaneous controls were defined as patients with hypertension whose primary care visit fell within the KnowCKD enrollment window (August 2025 to March 2026) and who did not participate in KnowCKD. Propensity scores were estimated by logistic regression on the variables shown above, and 1:2 nearest-neighbor matching was performed with a caliper of 0.2 × SD of the logit propensity score. Four KnowCKD participants could not be matched within the caliper and were excluded from the matched cohort. The standardized mean difference (SMD) before matching compares all KnowCKD participants (n=57) to all contemporaneous controls (n=182); the SMD after matching compares the matched KnowCKD (n=53) to matched controls (n=101). |SMD| < 0.10 indicates adequate balance.
Abbreviations: ACEi, angiotensin-converting enzyme inhibitor; ARB, angiotensin receptor blocker; BP, blood pressure; eGFR, estimated glomerular filtration rate; IQR, interquartile range; PCP, primary care provider; PSM, propensity score matching; SD, standard deviation; SMD, standardized mean difference.
Characteristics in this table were derived from the electronic health record and were used for the comparison between KnowCKD participants and contemporaneous controls and for propensity score estimation and matching. EHR-derived age and race variables may differ from the self-reported demographic characteristics presented in Table 2 because they were obtained from different data sources and coding structures.

Of the 57 KnowCKD participants, 53 (93.0%) were successfully matched to 101 contemporaneous controls. Matching improved covariate balance, with absolute SMDs below 0.10 for age, sex, primary care visit frequency, systolic blood pressure, eGFR, number of antihypertensive medication classes, and angiotensin-converting enzyme inhibitor or angiotensin receptor blocker use. A residual imbalance remained for Black race (SMD, −0.154).

In the propensity score-matched cohort, 20 of 53 KnowCKD participants (37.7%) completed albuminuria or proteinuria testing, compared with 13 of 101 matched controls (12.9%). This corresponded to a risk ratio of 2.93 (95% CI, 1.59–5.42; P<0.001) and an absolute risk difference of 24.9 percentage points (95% CI, 10.3–39.5).

In the sensitivity analysis, after excluding 2 KnowCKD participants and 18 controls with prior albuminuria testing, 21 of 55 KnowCKD participants (38.2%) and 19 of 164 controls (11.6%) completed testing (risk ratio, 3.30; 95% CI, 1.92–5.66).Results were similar when these exclusions were applied to the original propensity-matched cohort: 20 of 52 KnowCKD participants (38.5%) and 12 of 93 controls (12.9%; risk ratio, 2.98; 95% CI, 1.59–5.60).

### Participant Experiences With KnowCKD

Eight participants enrolled during 2026 participated in the post-intervention focus group. Three themes were identified: (1) KnowCKD increased awareness of albuminuria testing and prompted participants to raise testing during primary care visits; (2) participants considered the intervention acceptable and appropriate but recommended a clearer key message and less medical terminology; and (3) broader implementation would require a trusted means of health-system communication.

### KnowCKD increased awareness of albuminuria testing and prompted discussion during primary care visits

Participants described urine albumin testing as distinct from tests they routinely associated with primary care. KnowCKD increased their awareness that an additional urine test could be used to assess kidney health. Messages delivered close to scheduled visits served as reminders to discuss testing with their clinicians. One participant described the day-of-visit message as *“helpful for me to remember to ask her about the test,”* after which testing was ordered and completed that same day.

Importantly, participants described how the intervention prompted action even when detailed recall of its educational content was limited. As one participant explained, *“I also don’t remember word for word what was said, [but] it was enough to at least trigger that conversation for me to have with my primary.”* Participants described variation in clinician responses, including encounters in which testing was readily ordered and others in which testing did not occur.

### Participants found the intervention acceptable and appropriate but recommended a clearer key message and less medical terminology

Participants generally considered the amount and frequency of communication appropriate. One described the materials as *“right to the point,”* while another considered them sufficient to attract attention without being overwhelming. Participants valued the straightforward language and visual presentation of the infographic but recommended emphasizing the principal action—obtaining a urine test to detect early kidney disease—and reducing repetitive information. Several participants identified *albuminuria* as difficult to pronounce or remember. One commented, *“I’m not sure I can pronounce albuminuria,”* while another questioned whether unfamiliar terminology might reduce retention of the message.

### Participants emphasized the importance of trusted, contextualized, and actionable communication for broader implementation

Participants distinguished receiving KnowCKD as part of a research study from receiving similar messages during routine care. They emphasized that messages delivered outside the research setting would need to clearly identify their source and explain why the individual patient was receiving them. Several preferred established health-system communication channels, including an EHR patient portal. One participant stated, *“if it comes through MyChart, I’m really gonna pay attention to it because I know that’s specific to me.”*

Participants also favored messages explicitly linked to their health conditions and upcoming appointments rather than general health reminders. Finally, they identified practical barriers that could prevent testing after an order had been placed, including uncertainty about fasting, hydration before urine collection, and laboratory logistics. They recommended incorporating clear instructions to help patients complete the recommended testing.

## Discussion

In this mixed-methods study, KnowCKD, a brief behaviorally informed patient-facing intervention delivered before primary care visits, was associated with increased CKD awareness and knowledge, frequent patient-initiated discussions about urine testing for CKD diagnosis, and substantially greater completion of albuminuria or proteinuria testing among adults with hypertension. Qualitative findings showed that participants generally found the intervention clear, appropriately timed, and actionable, while identifying opportunities to simplify terminology, strengthen the central message, and improve delivery through trusted health-system channels.

These findings are particularly relevant in the context of the 2025 ACC/AHA multi-society hypertension guideline, which includes a Class 1 recommendation for UACR measurement in all adults with hypertension.^3^ Rapid uptake of this recommendation is important because detection of albuminuria has implications extending beyond CKD diagnosis. Albuminuria identifies patients at increased kidney and cardiovascular risk, improves risk stratification, and may identify individuals who benefit from therapies that reduce cardiovascular and kidney events.^20^ Yet albuminuria testing remains markedly underused among adults with hypertension, particularly among those without diabetes.^5^

Prior efforts to improve CKD-related testing and follow-up illustrate both the potential and the limitations of behavioral interventions. In a large pragmatic randomized trial in Japan, Fukuma and colleagues evaluated a behavioral economics–informed intervention among individuals identified with CKD through health screening.^21^ Although outreach increased subsequent physician visits compared with usual notification, adding behavioral-economics elements to the informational intervention did not further improve adherence or early health outcomes. The contrast with KnowCKD may reflect differences in both the target behavior and the timing of the intervention. KnowCKD was linked to an already scheduled primary care encounter and provided patients with a specific action to raise during that encounter, thereby reducing the need to initiate a separate healthcare visit. In addition, its content was iteratively developed with patients and paired education with a visit-proximate prompt.

Other patient-facing strategies have focused on reducing barriers to test completion itself. In a recent study of smartphone-enabled home UACR testing, providing patients with the ability to complete albuminuria testing at home substantially increased screening compared with usual care.^22^ This approach addresses a different point in the care pathway than KnowCKD: home testing reduces the logistical burden of completing a recommended test, whereas KnowCKD seeks to increase awareness, initiate discussion, and promote ordering during a clinical encounter. Together, these studies suggest that improving albuminuria testing may require interventions targeted at multiple steps in the testing pathway, including recognition of the need for testing, clinician ordering, and the practical completion of testing. Clinician-facing strategies such as electronic reminders and clinical decision support may further complement these patient-facing approaches by supporting recognition and action from the clinician side.

The qualitative findings provide additional insight into the features of KnowCKD that participants considered useful. Participants described urine albumin testing as distinct from laboratory testing they routinely associated with primary care and reported that the appointment-linked messages helped place testing on the agenda of their upcoming visit. Some participants recalled relatively little of the detailed educational content but nevertheless remembered that there was a kidney test to discuss with their clinician. Participants generally considered the amount and frequency of messaging appropriate but recommended a clearer central message and less reliance on terminology such as *albuminuria*, which several found difficult to pronounce or remember. They also emphasized that communication delivered outside a research setting should come from a recognizable health-system source, clearly explain why the patient is receiving it, and remain linked to an upcoming clinical encounter. These findings identify specific refinements that could improve KnowCKD before implementation in routine care.

The difference between patient-reported clinician ordering and EHR-confirmed test completion is also informative. Although 65% of participants reported that their clinician ordered a urine test, 37% had EHR-confirmed UACR or urine protein-to-creatinine ratio testing during observed follow-up. Several factors may explain this difference. Patient-reported ordering is subject to recall and reporting error, and some participants may have interpreted other urine studies, including routine urinalysis, as the test discussed during the intervention even though these studies did not meet the prespecified outcome definition. In addition, some qualifying albuminuria or proteinuria testing may have been completed outside the medical center and therefore not captured in the UCSF EHR. Ordering and completion also represent distinct steps in the care pathway: even when an appropriate test is ordered, completion requires the patient to present to a laboratory and provide a specimen. Participants in the qualitative evaluation described practical barriers, including uncertainty about fasting, hydration, and laboratory logistics.

Differences in follow-up time may also have contributed. The minimum follow-up among KnowCKD participants was 55 days, and 95% of observed post-intervention testing occurred within 60 days of the index date, suggesting that most testing occurred relatively soon after exposure. Nevertheless, later completion among participants enrolled near the end of the study may not have been captured. The observed EHR testing rate may therefore underestimate eventual test completion. Taken together, these findings suggest that interventions seeking to improve albuminuria testing should address the full sequence from awareness and patient engagement through clinician ordering and test completion.

KnowCKD was designed as a brief and reproducible intervention, and its structure lends itself to integration into routine clinical workflows. Eligibility can be determined from structured EHR information, delivery can be linked automatically to an upcoming primary care appointment, and messages can be sent through existing patient communication systems. Such an approach could minimize additional clinician burden while reaching patients at a time when they can act on the information. However, participants in the present study actively enrolled in research and knew they were receiving a study intervention. Engagement with automated outreach delivered as part of routine care may differ. The next step should therefore be a rigorous pragmatic randomized evaluation embedded directly within the health system, in which eligible patients are identified from the EHR and randomized to receive appointment-linked KnowCKD outreach or usual care without traditional participant enrollment before intervention delivery, when ethically and operationally permissible. Such a design would provide a more direct estimate of effectiveness, reach, and scalability under real-world conditions and allow evaluation across more diverse populations and clinical settings.

Several limitations should be considered. First, KnowCKD participation was not randomized, and individuals who chose to enroll may have differed from contemporaneous controls in characteristics associated with their likelihood of completing testing. Propensity score matching improved balance across measured characteristics and yielded estimates similar to those of the primary analysis, but residual and unmeasured confounding cannot be excluded. Second, the study was conducted within a single academic health system and included a relatively small number of KnowCKD participants, which may limit generalizability. Third, observation time differed between intervention participants and controls because controls had no analogous intervention date. The primary approach provided controls with a longer opportunity for testing than the median follow-up among KnowCKD participants but differential follow-up remains a design limitation. Fourth, survey outcomes were self-reported and may be affected by recall or reporting bias. Fifth, the qualitative evaluation consisted of a single focus group restricted to participants enrolled during 2026 to minimize recall bias and was intended to characterize experiences and implementation considerations rather than estimate the prevalence of specific perspectives. Qualitative coding was performed by a single investigator, which may have increased susceptibility to interpretive bias.

In conclusion, KnowCKD was associated with increased CKD awareness and knowledge, frequent patient-initiated discussions about kidney testing, and substantially greater EHR-confirmed albuminuria or proteinuria testing among adults with hypertension. As health systems seek to implement the 2025 ACC/AHA multi-society recommendation for routine albuminuria assessment in hypertension, behaviorally informed patient activation may provide a scalable complement to clinician-facing and test-completion strategies. Pragmatic randomized evaluation within routine health-system workflows will be needed to determine whether these findings can be reproduced at scale and across more diverse clinical settings.

## Data Availability

All data produced in the present study are available upon reasonable request to the authors

